# Meta-analysis of RNA sequencing data from 534 skin samples shows substantial IL-17 effects in non-lesional psoriatic skin

**DOI:** 10.1101/2023.11.03.23298021

**Authors:** Åshild Øksnevad Solvin, Konika Chawla, Marita Jenssen, Lene Christin Olsen, Anne-Sofie Furberg, Kjersti Danielsen, Marit Saunes, Kristian Hveem, Pål Sætrom, Mari Løset

**Author notes:** **Correspondence: HUNT Center for Molecular and Clinical Epidemiology**, Department of Public Health and Nursing, Norwegian University of Science and Technology, NTNU Håkon Jarls gt. 11, 7491 Trondheim, Norway, /. These authors contributed equally to this work (Åshild Øksnevad Solvin, Konika Chawla). These authors jointly supervised this work (Pål Sætrom, Mari Løset).

## Abstract

Psoriasis is a common chronic inflammatory skin disease characterized by disturbed interactions between infiltrating immune cells and keratinocytes. To enhance our understanding of the underlying molecular and cellular mechanisms driving psoriasis pathobiology, and to identify potential biomarkers for disease severity, we conducted RNA sequencing of skin biopsies from 75 patients with psoriasis vulgaris and 46 non-psoriatic controls. To increase the robustness of the results, we meta-analysed our data with four publicly available datasets, bringing the total number of samples to 534. By comparing lesional psoriatic (PP) to healthy control (NN) skin, we identified 2269 differentially expressed genes (DEGs) (|log_2_FC|>1.0, FDR <0.1), and 58 DEGs when comparing non-lesional psoriatic (PN) to NN skin. We also identified 54 DEGs associated with disease severity (PASI ≥10 vs. PASI <10). Cellular deconvolution analysis showed that differentiated keratinocytes emerged as the most prominent cell type among the DEGs in PP/NN. Functional enrichment analysis in PN/NN revealed several IL-17 related pathways and confirmed a previously reported ‘pre-inflammatory signature’ across all psoriatic skin. This study provides insights into the psoriasis transcriptome and identifies a severity-specific signature, which may serve as candidate for future studies aimed at identifying psoriasis biomarkers and predicting disease progression.

## INTRODUCTION

Psoriasis is a chronic inflammatory skin disease affecting 1-2% of the global population. ^1^ The prevalence in Norway is among the highest in the world, ranging from 5.8-11.4%. ^2,3^ Psoriasis arises from a complex interplay between genomics and environmental influences, with an estimated heritability of >60%.^4,5^ Psoriasis vulgaris, the most common subtype, is characterized by well-demarked, erythematous, scaly plaques. Cellular changes in the affected skin involve infiltration of immune cells, hyperproliferation of epidermal keratinocytes and vascular hyperplasia.

Much of our knowledge of the molecular contributors of psoriasis stems from gene expression (RNA) data from psoriatic skin samples. ^6,7^ Over the last two decades, RNA sequencing (RNA-seq) has revolutionized the opportunities to perform hypothesis-free global gene expression profiling. Results from such studies have shed light on disease mechanisms and pathology, ^6–8^ and established psoriasis as an interleukin (IL)-23/T helper (Th)-17 dominated disease. Previous studies have identified differentially expressed genes (DEGs) and perturbed pathways involved in innate and adaptive immunity, as well as in keratinocyte differentiation. ^7,9^ RNA-seq has also shed light on the regulatory roles of long non-coding RNAs (lncRNA) in psoriasis including the discovery of Psoriasis susceptibility-related RNA gene induced by stress (*PRINS*). ^10^ While much of our knowledge of psoriasis pathobiology is derived from gene expression data, previous studies have included limited numbers of non-lesional psoriatic samples (≤27), with only few studies exploring the impact of disease severity on gene expression. Moreover, a comprehensive profiling of both polyadenylated and non-polyadenylated lncRNAs in a large psoriatic sample has been lacking.

In this study, we explore the transcriptional profile of psoriasis by integrating RNA-seq data from a locally curated dataset and four publicly available datasets. We identify the contribution of different cell types through deconvolution analyses. Additionally, we assess whether the RNA expression differs between individuals with mild versus moderate to severe disease, thereby shedding light on potential biomarkers for disease progression and treatment.

## METHODS

### Human samples, RNA extraction and sequencing

The collection of samples and methods used for this study has been fully described previously. ^11^ All participants had provided written informed consent. The study was approved by the Regional Committee for Medical and Health Research Ethics in Mid-Norway (2016/281) and North-Norway (2016/789) and adheres to the Declaration of Helsinki Principles. In brief, 4 mm punch biopsies were collected from lesional (PP), non-lesional (PN) and healthy control (NN) skin, snap frozen in liquid nitrogen and stored at –80 °C. Psoriasis severity was assessed by Psoriasis Area and Severity Index (PASI). Total RNA was extracted and purified using the mirVana Isolation Kit (Applied Biosystems, Foster City, CA, USA) according to the manufacturer’s instructions. RNA integrity number (RIN) was assessed using the Agilent RNA 6000 Nano kit on a 2100 Bioanalyzer chip (Agilent Technologies, Inc., Santa Clara, CA, USA). The effect of RIN value was assessed by a multidimensional scaling plot (Supplementary figure 1). All samples were included in further analyses as the samples with lower RIN did not separate from the remaining samples. RNA sequencing libraries were generated using SENSE mRNA-Seq library prep kit according to manufacturer’s instructions (Lexogen GmbH, Vienna, Austria). Single-end read sequencing was performed on an Illumina HiSeq4000 instrument (Illumina, Inc., San Diego, CA, USA). Low-quality reads were filtered out using fastq_quality_filter. The remaining high-quality reads were then aligned to the human reference genome GRCh38.92 using STAR v.2.4.2a, ^12^ and featureCounts ^13^ was used to annotate the reads to distinct genes in GRCh38.92. Genes with an average count per million of less than 0.1 across all samples were filtered out. This gave us a normalized count matrix of 22 530 expressed genes. For the purpose of this study, genes were classified into four groups (protein coding, long non-coding, pseudogene and ‘other’) based on Gene/Transcript biotypes as defined by GENCODE and Ensembl (Supplementary table 1).

### Differential expression analysis

A linear model with coefficients for sample type (PP, PN NN) and adjusted for sex, age and BMI was used to identify DEGs. We did three differential expression analysis (PP vs. NN; PP vs. PN; and PN vs. NN). The count matrix was prepared with number of reads mapping to each gene per sample. It was read as a DGEList in R using the egdeR ^14^ and later transformed by the voom algorithm from the limma package of Bioconductor. ^15,16^ Fold change (FC) for each gene was calculated across the three comparisons and then log2 transformed. The Benjamini-Hochberg method was used to control the false discovery rate (FDR). We used |log_2_FC|>1.0 and FDR <0.1 to declare significance. To determine if any mRNAs were related to psoriasis severity, cases were further separated into PASI <10 and PASI ≥10 before differential expression analysis. To enhance the robustness of the identified DEGs, we reanalysed data from four publicly available datasets (NCBI GEO accession numbers: SuperSeries GSE63980 (GSE54456 and GSE63979), GSE121212 and GSE74697) and combined the count matrices from these datasets with the local dataset. This resulted in a normalized count matrix of 17 662 expressed transcripts. We applied the same statistical method to this combined count matrix to identify a meta-analysed set of DEGs. Note that for the meta-analysis, the linear model included sample type, but with study Id as the only adjustment, as information on sex, age and BMI was only available from one study (GSE21212). In addition, we analysed each dataset individually to identify consistencies and irregularities in DEGs between the studies.

### Deconvolution analysis

To estimate cell fractions from whole tissue samples, we used CIBERSORTx. This is an analytical tool tailored for digital cytometry. This tool is designed to estimate cell type abundance in whole tissue RNA sequencing data, without the need for physical dissociation. ^17^ The method utilizes data from single-cell sequenced samples to create a cell type signature matrix. This matrix is subsequently applied to bulk tissue RNA profiles to infer cell type proportions. ^18^ In our analysis, we imported DEGs from the meta-analysis and utilized healthy skin data, as presented by Reynolds et al. ^19^ as the signature matrix.

### Functional enrichment analysis

To predict biological effects of the dysregulated RNAome in psoriasis, we performed enrichment analysis by importing DEGs to QIAGEN Ingenuity Pathway Analysis (IPA) (QIAGEN Inc., https://digitalinsights.qiagen.com/IPA). ^20^ The gene identifiers of the transcripts were mapped to their respective gene objects in the Ingenuity Pathway Knowledge Base. IPA was used to bioinformatically identify canonical pathways associated with the DEGs.

## RESULTS

The local dataset included 75 psoriatic cases in total (71 PP, 74 PN) and 46 non-psoriatic controls (46 NN). In the meta-analysis, sample size was increased to 534 samples (216 PP, 128 PN, 190 NN) (Supplementary table 2).

### Differential expression analysis

Multidimensional scaling of the RNA expression profiles in the locally collected samples showed a distinct separation of the PP samples from PN and NN samples, with the latter two overlapping (Figure 1a). For the meta-analysis, differences between the datasets constituted the primary and secondary source of variation (Dim1 and Dim2). There was a tendency towards separation between sample groups PP, PN and NN (Figure 1b). Correlations between DEGs in the local dataset and those in the meta-analysis were strong (Spearman correlation coefficient: 0.86), as were correlations with the Michigan datasets (Spearman correlation coefficient: 0.71), and the Kiel dataset (Spearman correlation coefficient: 0.77) (Figure 1c). A higher correlation was observed between mRNAs identified as differentially expressed in all datasets (dark green) compared to those differentially expressed in fewer datasets (light green/turquoise).

**Figure 1.**
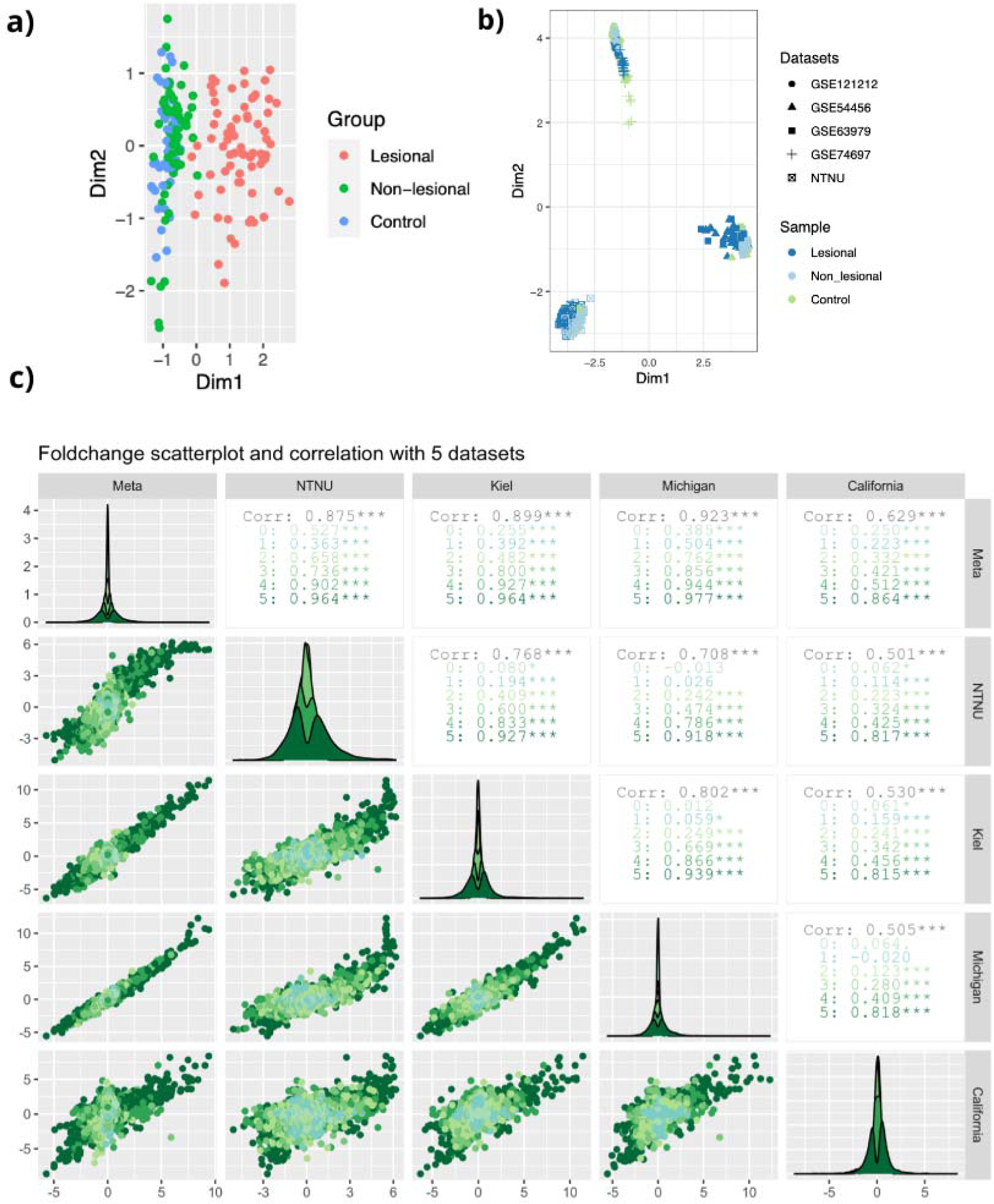
Gene expression analysis in the local dataset and meta-analysis datasets. a) Multidimensional scaling (MDS) plot showing lesional (PP), non-lesional (PN) and control (NN) samples for the local dataset. b) MDS plot for the meta-analysis dataset, including sample source. c) Correlogram presenting scatterplots and Spearman’s rank correlation coefficient across the datasets. The numbers 0 to 5 denote the number of datasets in which a differentially expressed gene (DEG) was found to be significant.

In the local dataset, we identified 4992, 4708 and 29 DEGs (|log_2_ FC| ≥1 and FDR ≤0.1) in the PP/NN, PP/PN and PN/NN comparisons, respectively (Supplementary table 3). The meta-analysis identified 2269, 2019 and 58 DEGs in the PP/NN, PP/PN and PN/NN comparisons, respectively. In PP/NN, 1849 DEGs (34.2%) were shared between the local dataset and the meta-analysis (Figure 2a). In PN/NN, only 3 DEGs (3.6%) were identified as common between the local dataset and the meta-analysis (Figure 2b).

**Figure 2.**
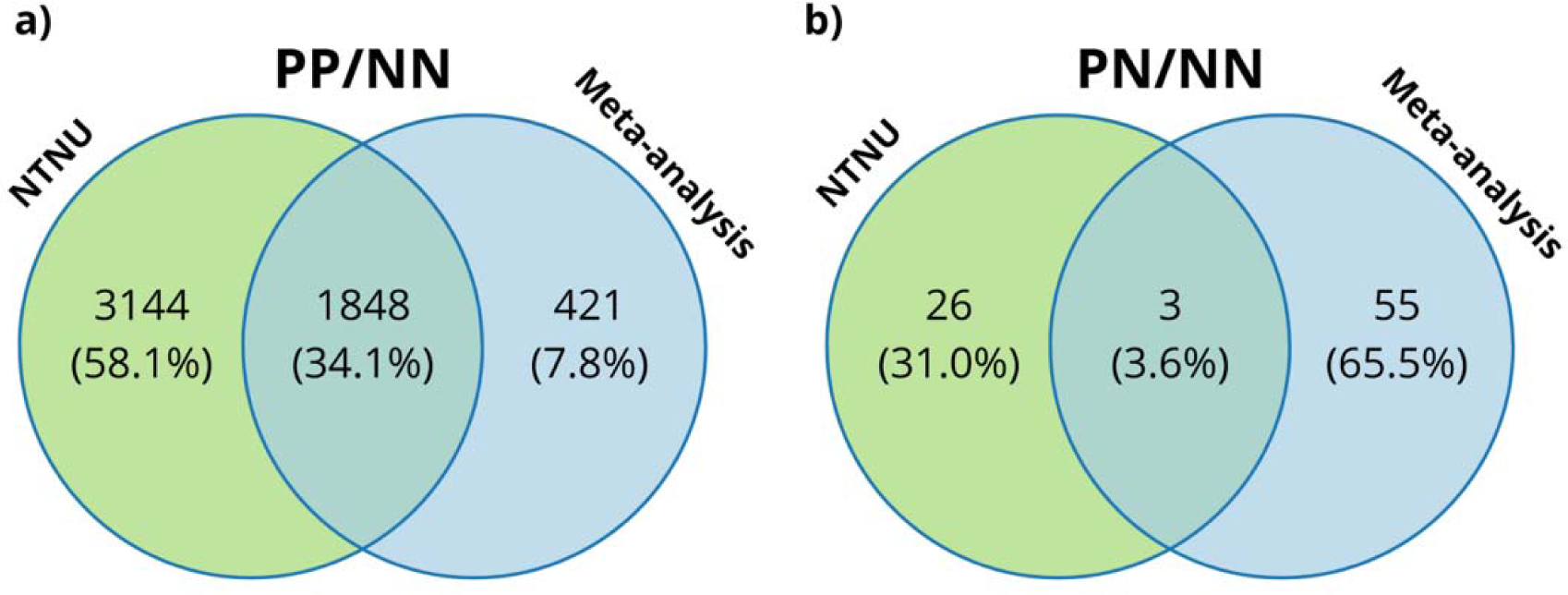
Venn diagrams illustrating common differentially expressed genes (DEGs) between the local dataset and the meta-analysed dataset for a) PP/NN and b) PN/NN comparisons.

Using Ensemble as our reference, we classified the identified transcripts into four distinct RNA biotypes: (i) protein-coding, (ii) long non-coding RNA, (iii) pseudogene, and (iv) ‘other’. The majority of the DEGs identified (ranging from 86.2%-88.0%) fell into the protein-coding category (Supplementary table 4). The top 10 protein-coding DEGs in the PP/NN and PN/NN comparisons, sorted by absolute log fold change can be found in Table 1a and 1b. Lists of the top 10 DEGs in the other biotypes (long non-coding, pseudogene, other) are provided in supplementary tables 5-6.

**Table 1.**
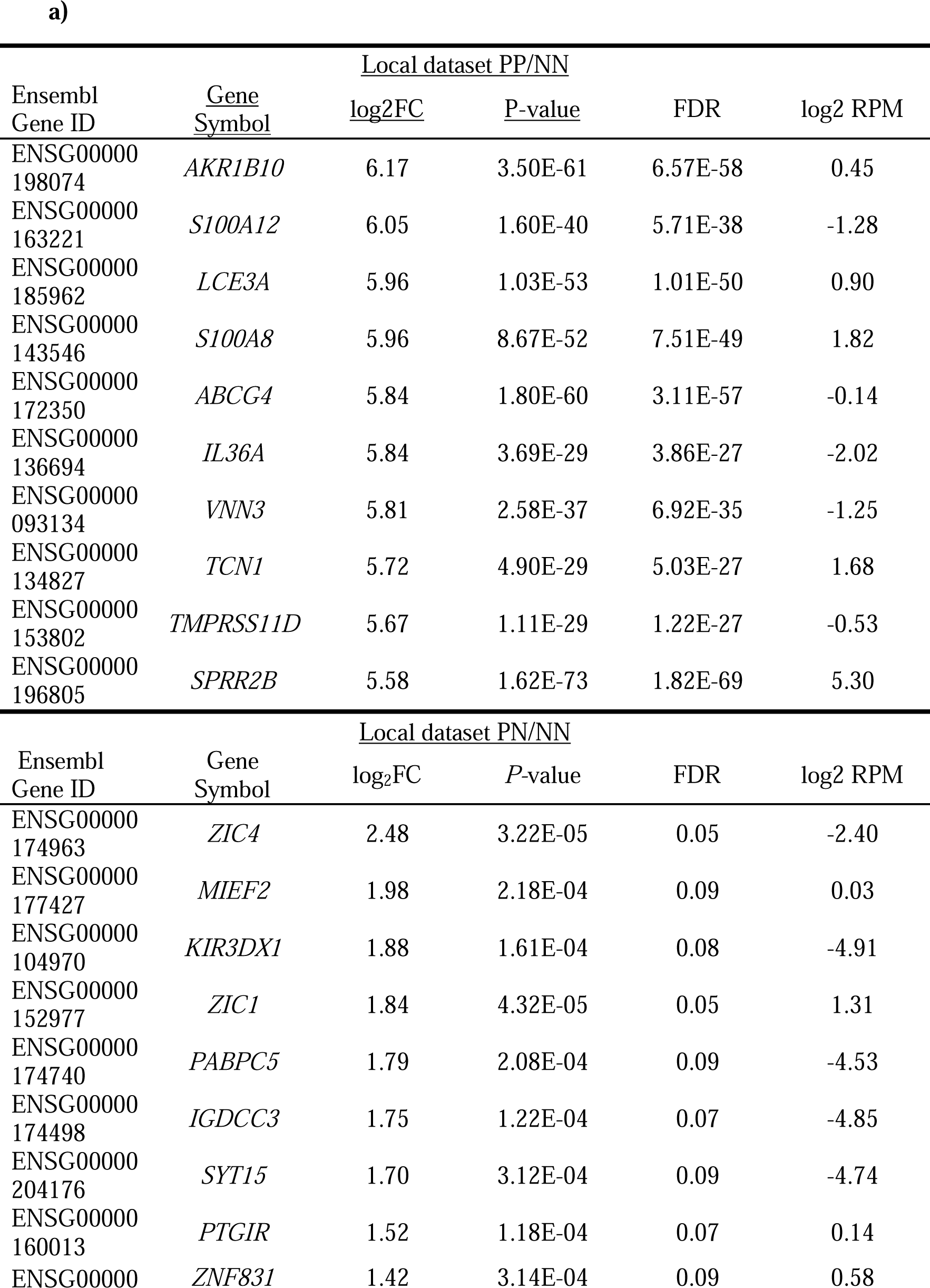
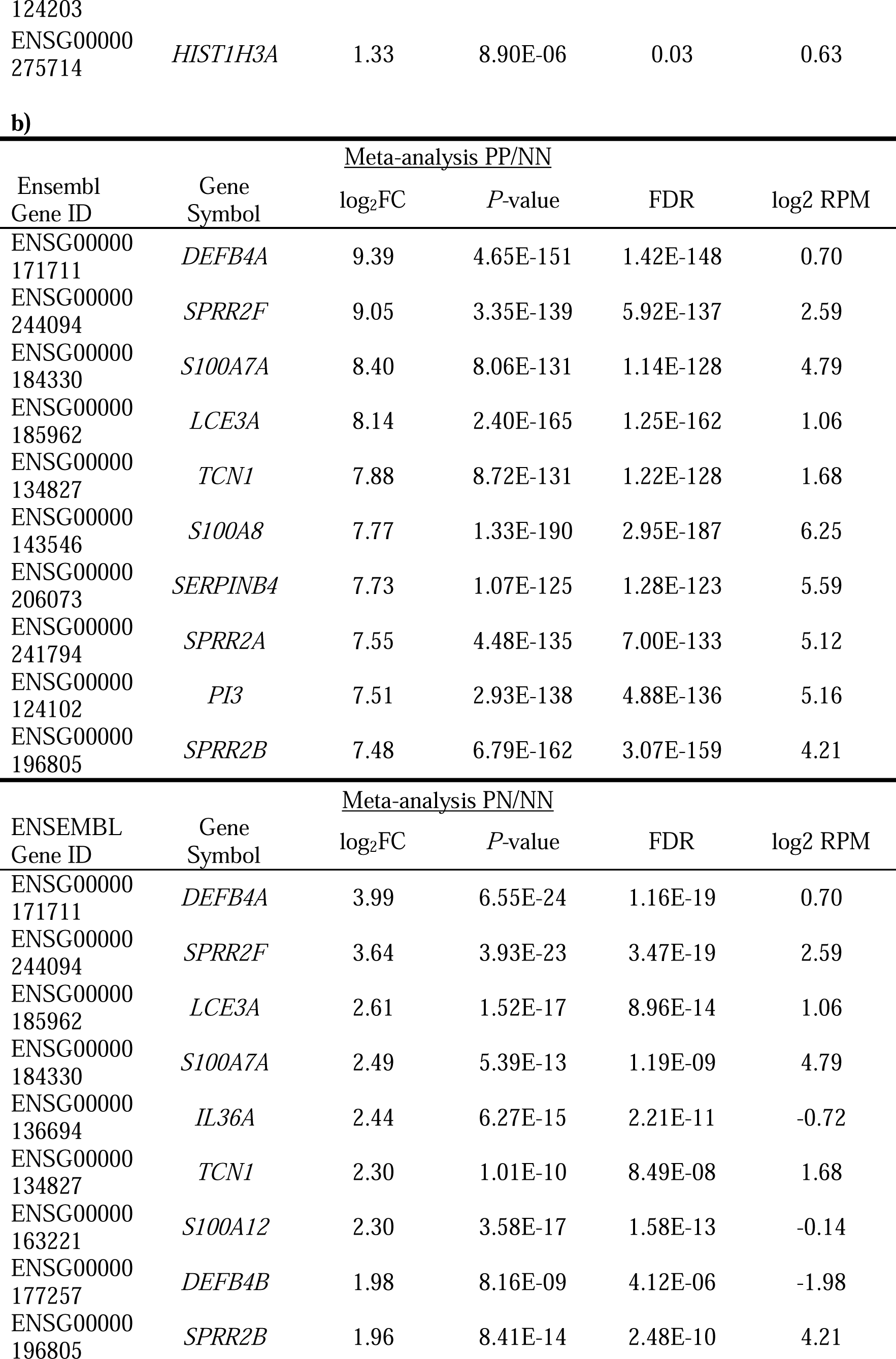
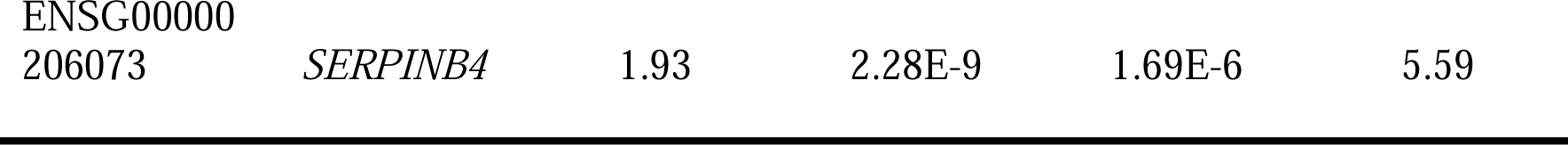
Top 10 differentially expressed protein-coding genes sorted by absolute fold change (|log_2_ FC|) in the PP/NN and PN/NN contrasts for a) the local dataset and b) the meta-analysis. The genes’ estimated difference in mean expression between the contrasts is presented as log2 reads per million (RPM)

We also compared PP in PASI ≥10 to PP in PASI <10, as well as PN in PASI ≥10 to PN in PASI <10. In PP skin, we identified 7 DEGs between PASI ≥10 and PASI <10 (FDR ≤0.1 and |log_2_ FC| ≥1). In PN skin, we identified 47 DEGs (FDR ≤0.1 and |log_2_ FC| ≥1) between the PASI ≥10 and PASI <10 groups. The top protein-coding DEGs in the severity-specific analysis, sorted by absolute log fold change, can be found in Table 2a and 2b. Lists of the remaining DEGs can be found in supplementary tables 7-8.

**Table 2.** Top severity-associated (PASI≥ 10 vs. PASI <10) protein coding DEGs sorted by absolute fold change (|log_2_ FC|) in PP and PN. The estimated difference in mean expression between the contrast is presented as log2 reads per million (RPM).

| a) |  | PP |  |  |  |
| --- | --- | --- | --- | --- | --- |
| Ensembl<br>Gene ID | Gene<br>Symbol | log<sub>2</sub>FC | P-value | FDR | log2 RPM |
| ENSG00000224689 | *ZNF812P* | 3.29 | 7.34E-07 | 0.01 | -4.41 |
| ENSG00000167483 | *FAM129C* | 3.08 | 3.07E-05 | 0.06 | -4.44 |
| ENSG00000177257 | *DEFB4B* | 2.95 | 2.47E-05 | 0.06 | -4.84 |
| ENSG00000167612 | *ANKRD33* | 2.61 | 2.06E-05 | 0.06 | -5.12 |
| ENSG00000172867 | *KRT2* | -1.92 | 1.22E-05 | 0.06 | 9.54 |
| ENSG00000162616 | *DNAJB4* | -1.86 | 1.80E-05 | 0.06 | 0.26 |
| ENSG00000104368 | *PLAT* | 1.24 | 2.72E-05 | 0.06 | 4.65 |
| b) |  | PN |  |  |  |
| Ensembl<br>Gene ID | Gene<br>Symbol | log<sub>2</sub>FC | P-value | FDR | log2 RPM |
| ENSG00000170374 | *SP7* | 3.55 | 1.17E-05 | 0.02 | -3.75 |
| ENSG00000148335 | *NTMT1* | -3.27 | 3.26E-07 | 0.01 | 1.25 |
| ENSG00000174498 | *IGDCC3* | 3.11 | 1.10E-06 | 0.01 | -4.85 |
| ENSG00000196353 | *CPNE4* | -2.98 | 1.16E-04 | 0.06 | -0.45 |
| ENSG00000171155 | *C1GALT1C1* | -2.86 | 3.97E-05 | 0.04 | -0.36 |
| ENSG00000186458 | *DEFB132* | 2.72 | 3.40E-04 | 0.10 | -3.61 |
| ENSG00000134153 | *EMC7* | -2.64 | -1.21 | 6.72E-06 | 0.02 |
| ENSG00000143001 | *TMEM61* | -2.69 | -2.21 | 2.24E-04 | 0.08 |
| ENSG00000271271 | *UGT2A2* | 2.69 | -4.36 | 1.25E-04 | 0.06 |
| ENSG00000204930 | *FAM221B* | 2.71 | -4.23 | 2.38E-04 | 0.08 |

### Differentiated KCs contribute to altered cell composition in PP/NN

Through cellular deconvolution of the DEGs in the PP/NN comparison, we found an increased fraction of differentiated keratinocytes and a decreased fraction of undifferentiated keratinocytes (Figure 3a). Additionally, there was a decreased fraction of various dermal non-immune cells, including fibroblasts F2 and pericytes. The proportion of antigen presenting cells differed between cell types. Specifically, we identified increased fractions of macrophages and dendritic cells D2, along with decreased fractions of dendritic cells D1, inflammatory macrophages and Langerhans cells in PP skin. Moreover, PP skin exhibited an elevated fraction of T helper cells and reduced fraction of innate lymphoid/natural killer cell compared to NN skin. When comparing PN to NN, fewer significant differences in cell type fractions were observed (Figure 3b). There was an increased fraction of melanocytes in PN skin compared to NN skin, while no significant differences were found in the fractions of other non-immune epidermal or dermal cell types. We also identified an increased fraction of innate lymphoid/natural killer cells and monocyte-derived dendritic cells, and a decreased fraction of dendritic cells 1 in PN skin. Among the antigen-presenting cells, only T helper cells exhibited a significant differential presence, showing an increased fraction in PN skin compared to NN skin.

**Figure 3.**
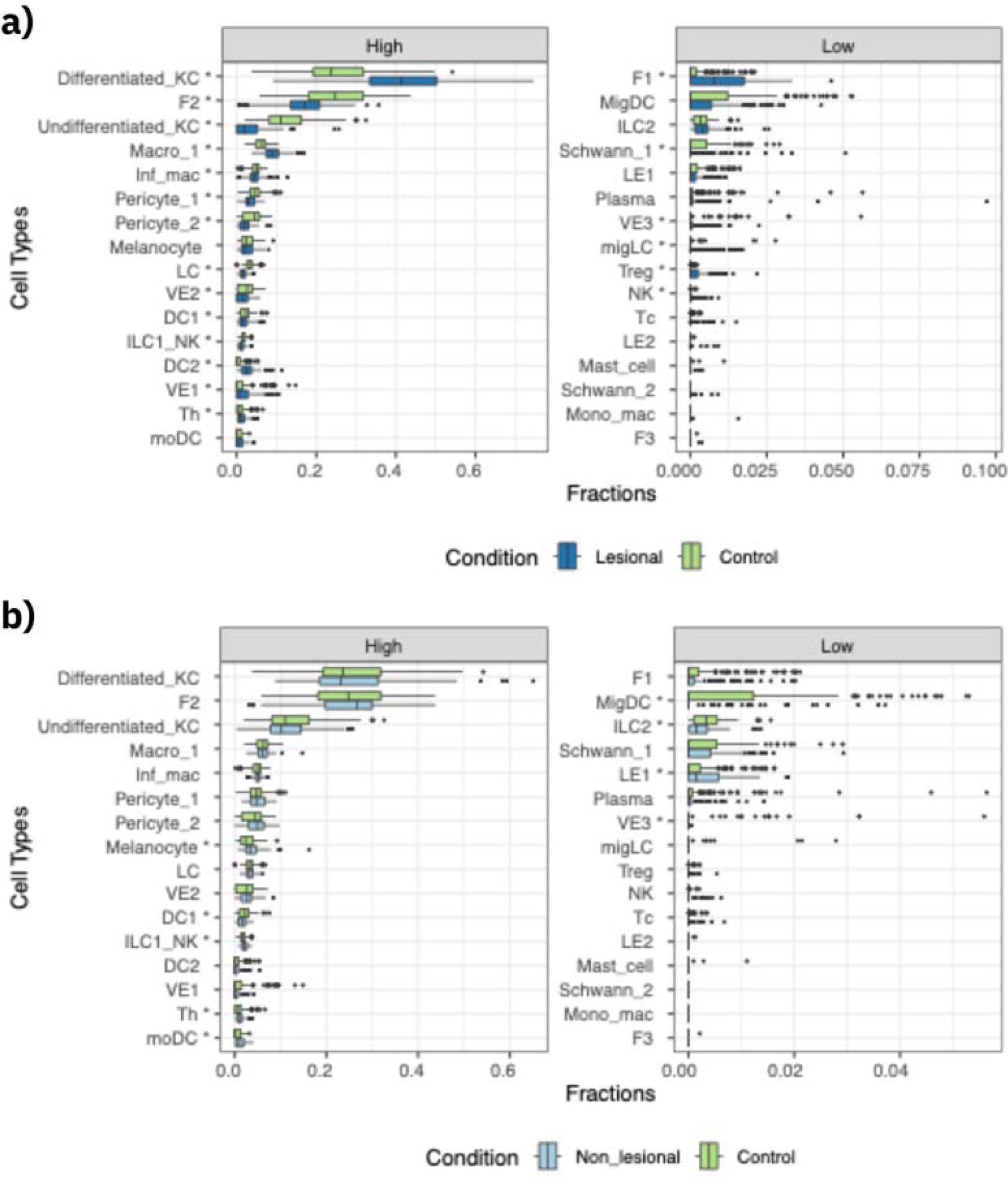
Box plot of deconvoluted cell type fractions estimated by CIBERSORTx for a) PP/NN and b) PN/NN comparisons. Cell types were ordered based on their mean fractions; a star (*) after the cell type name signifies a statistically significant difference between the sample types (adjusted p-value <0.05). Separate plots were created for cell types with high (left) and low (right) fractions to enhance readability. Abbreviations: KC, keratinocytes; F, fibroblast; macro/mac, macrophage; inf, inflammatory; LC, Langerhans cell; VE, vascular endothelium; DC, dendritic cell; ILC, innate lymphoid cell; NK, natural killer cell; Th, T helper cell; mo/mono, monocyte-derived; mig, migratory; LE, lymphatic endothelium; Treg, T regulatory T cell, Tc; cytotoxic T cell.

### Functional enrichment identified cytokine storm and IL-17 signaling as main pathways

For the DEGs unrelated to PASI, we applied the gene lists from the meta-analysis for functional enrichment. The results of the DEG analysis and functional enrichment were very consistent for PP/NN and PP/PN, consequently, we do not present further details for PP/PN. Within the PP/NN comparison, the top five canonical pathways identified were ‘Pathogen Induced Cytokine Storm Signaling Pathway’ (*P*-value: 3.14 × 10^−15^), ‘Agranulocyte Adhesion and Diapedesis’ (*P*-value: 3.70 × 10^−15^), ‘S100 Family Signaling Pathway’ (*P*-value: 6.25 × 10^−14^), ‘Granulocyte Adhesion and Diapedesis’ (*P*-value: 1.53 × 10^−13^), and ‘Atherosclerosis Signaling’ (*P*-value: 2.18 × 10^−12^) (Figure 4a, Supplementary table 9). The top predicted upstream regulators included TNF (*P*-value: 5.19 × 10^−58^), IFNG (*P*-value: 2.77 × 10^−56^) and beta-estradiol (*P*-value: 6.97 × 10^−49^) (Supplementary table 10).

**Figure 4.**
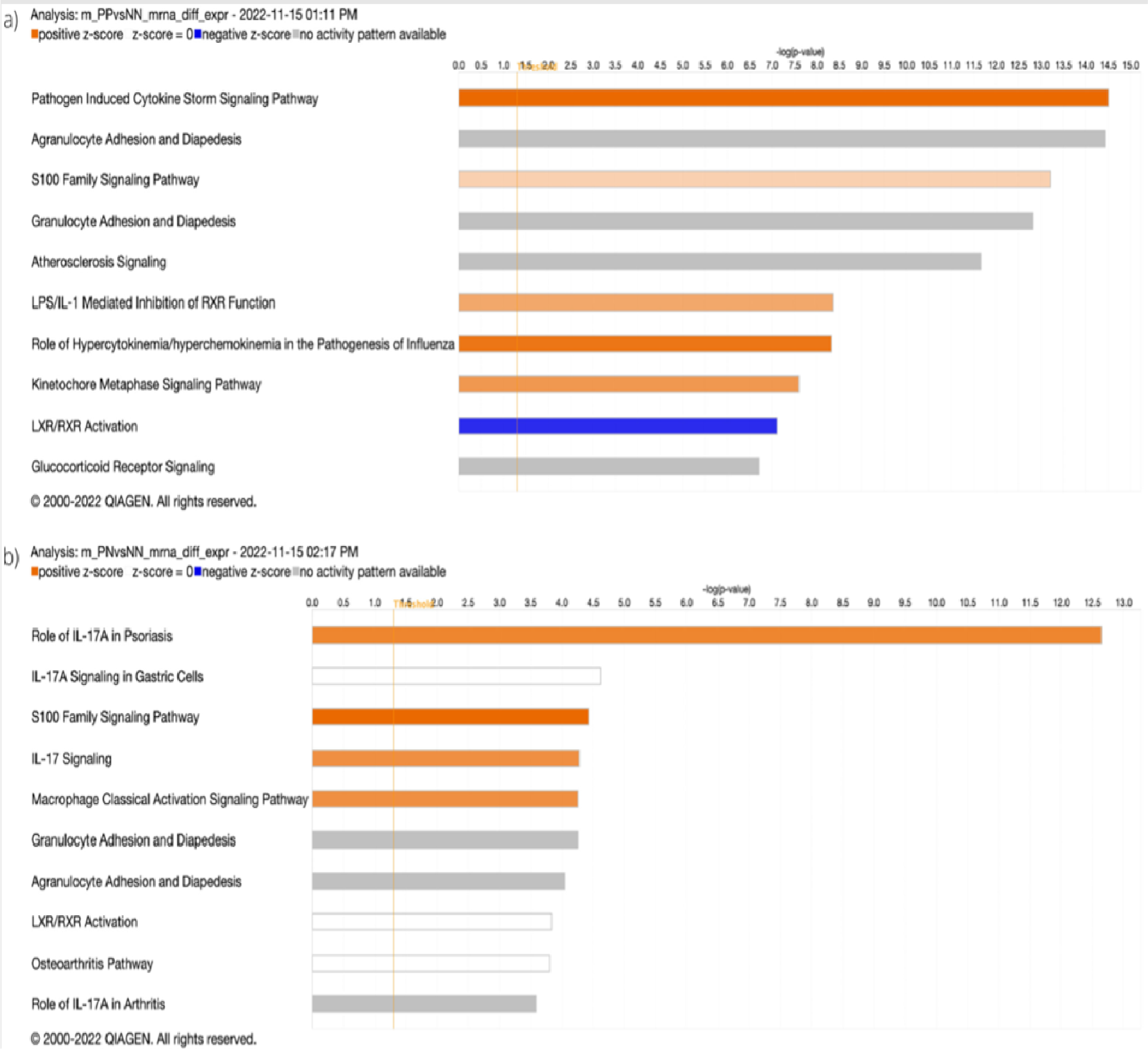
Top 10 enriched canonical pathways for the a) PP/NN and b) PN/NN comparisons in the meta-analysis. Orange, white and blue indicated increased, neutral and decreased pathway activity respectively. Grey indicated no activity pattern available.

In PN/NN, the top five canonical pathways were ‘Role of IL-17A in Psoriasis’ (*P*-value: 2.27 × 10^−13^), ‘IL-7A Signaling in Gastric Cells’ (*P*-value: 2.26 × 10^−5^), ‘S100 Family Signaling Pathway’(*P*-value: 3.73 × 10^−5^), ‘IL-17 Signaling’ (*P*-value: 5.27 × 10^−5^) and ‘Macrophage Classical Activation Signaling Pathway’ (*P*-value: 5.53 × 10^−5^) (Figure 4b, Supplementary table 11). The top predicted upstream regulators included IL-1A (*P*-value: 1.28 × 10^−16^), IL-22 (*P*-value: 1.92 × 10^−16^), and IL-17A (*P*-value: 4.69 × 10^−16^) (Supplementary table 12).

### Functional enrichment of severity associated DEGs

Enrichment analysis of DEGs, comparing PASI ≥10 to PASI <10 in PP skin, identified the following top canonical pathways: ‘Role of IL-17A in Psoriasis’ (*P*-value: 3.57 × 10^−3^), ‘Differential Regulation of Cytokine Production in Intestinal Epithelial Cells by IL-17A and IL-17F’ (*P*-value: 5.85 × 10^−3^), ‘Coagulation system’ (*P*-value: 8.90 × 10^−3^), ‘IL-17A Signaling in Airway Cells’ (*P*-value: 1.70 × 10^−2^), and ‘TREM1 Signaling’ (*P*-value: 1.95 × 10^−2^) (Supplementary table 13). The top upstream regulators included DACH1 (*P*-value: 5.89 × 10^−55^), ganglioside GM2 (*P*-value: 2.83 × 10^−4^) and proadifen (*P*-value: 2.83 × 10^−4^) (Supplementary table 14). In PN skin, the top canonical pathways were ‘IL-13 Signaling’ (*P*-value: 1.15 × 10^−2^), ‘alfa-tocopherol Degradation’ (*P*-value: 1.39 × 10^−2^), ‘Parkinson’s Signaling’ (*P*-value: 2.22 × 10^−2^), ‘Glutaryl-CoA Degradation (*P*-value: 2.63 × 10^−2^) ’ and ‘IL-17 Signaling’ (*P*-value: 2.82 × 10^−2^) (Supplementary table 15). The top upstream regulators included miR-629-5p (*P*-value: 1.42 × 10^−3^) and N-carbamyl-L-glutamate (*P*-value: 1.42 × 10^−3^) (Supplementary table 16).

## DISCUSSION

In this study, we identify a substantial number of DEGs in psoriasis most of which were protein–coding, and a severity-specific gene signature for both PP and PN skin. The cell-specific analysis highlights an elevated fraction of differentiated KCs in psoriasis. Functional enrichment analyses underscore significant IL-17 effects in non-lesional psoriatic skin, providing support for a ‘pre-inflammatory signature’. ^21,22^

Meta-analysis of 534 skin biopsies identified a total of 2269 DEGs in PP/NN and 58 DEGs in PN/NN. DEGs in both PP/NN and PN/NN indicated an increased representation of S100 signaling processes. The S100 family of proteins play diverse roles and specific members have been implicated in psoriasis development. *S100A7* (also known as psoriasin) exhibits upregulation in both skin and sera of individuals with psoriasis. ^23^ In PP/NN, psoriasis autoantigens *KRT17* and *PLA2G4D* were upregulated (logFC 1.89 and 4.02, respectively). Like a previous publication, ^21^ we found similar expression levels of IL17A in PN/NN. However, the top canonical pathways for the meta-analysis in PN/NN indicate significant IL-17 effects, corroborating results reported by Keermann et al.. ^9^ In fact, three out of the top five most enriched pathways were all related to IL-17 signaling pathways. Moreover, we replicated increased expression of *DEFB4* and *S100A7*, established as known IL-17 signature genes. ^24^

Our results encompass both poly-A and non-poly-A lncRNAs. In PP/NN, we observed upregulation of *KLHDC7B-DT* (log_2_FC: 4.35) a lncRNA experimentally shown to contribute to keratinocyte hyperproliferation and skin inflammation in psoriatic keratinocytes through activation of STAT3 and JNK signalling pathways. ^25^ Additionally, the upregulation of *UCA1* (log_2_FC: 1.97), a keratinocyte-specific lncRNA, underscores its role in the interplay between psoriatic keratinocytes and immunity. ^26^ While *PRINS* has been previously implicated in psoriasis, ^10^ our study did not replicate this. Given their finely tuned regulatory capacities, lncRNAs exhibit great potential as biomarkers for diagnosis, disease development and treatment in psoriasis. ^27^ Hence, uncovering disease-associated lncRNAs in psoriasis may pave the way for novel biomarker discovery.

Deconvolution analysis using CIBERSORTx unveiled an increased proportion of differentiated keratinocytes in PP skin and conversely, a decreased proportion of undifferentiated keratinocytes in PP skin compared to NN skin. These findings align with the work of Reynolds et al., ^19^ emphasizing the rapid turnover of keratinocytes in psoriatic skin. Furthermore, our analysis identified an elevated fraction of inflammatory Langerhans cells and macrophage 1 in PP/NN skin. However, disparities emerged between our results and those presented by Reynolds et al. Notably, we did not replicate increased fractions of cytotoxic T cells or macrophage 2. Direct comparison is challenging due to methodological variations, including differences in reported cell categories within single cell experiments.^19,28^ Moreover, our utilization of healthy skin as the signature matrix, rather than the psoriatic skin, may also influence interpretation of these results.

We identified a severity-specific signature that may offer promising candidates for future investigations in predictive biomarkers for severe disease. *FAM129C*, suggested as a transcriptional marker of plasmacytoid dendritic cells (pDCs) ^29^ was the most dysregulated (log_2_FC 3.08) in PP skin with PASI ≥10. pDCs have been recognized in the development of psoriasis ^30^ and our results lends further weight to their roles increased disease activity. *DEFB4B,* a skin antimicrobial peptide, was also upregulated (log_2_FC: 2.95), in line with a previous study finding that this gene is downregulated in psoriatic skin successfully treated with dithranol. ^31^ It is also upregulated in the chronic inflammatory skin disease hidradenitis suppurativa. ^32^ In PN skin, *DEFB132*, an antimicrobial peptide was upregulated. The DEGs in PP PASI >10 were largely enriched for IL-17 related pathways. IL-17 concentration in scales correlates positively with PASI in psoriatic males. ^33^ IL-17 activity has previously been identified as a psoriasis-defining transcriptomic feature ^34^ and our results point to an association between IL-17 activity and increasing psoriasis severity. Kim ^35^ and Choudhary ^36^, identified IL-17 responses as a feature of mild psoriasis. However, these studies compared PP to NN skin and results are therefore not directly comparable. Additionally, in our analysis, the activity pattern (activated or repressed) could not be determined and is based on a relatively low number of DEGs. Pathways including ‘coagulation system’ and ’ triggering receptor expressed on myeloid cells-1 (TREM1)-signaling’ were also enriched in the PP skin of individuals with PASI ≥10. The interplay between the coagulation and inflammatory systems has been recognized ^37^ and abnormal platelet activation has been demonstrated in psoriasis. ^38^ While TREM-1 expression increases during inflammation and its involvement in psoriasis pathogenesis has been suggested, ^39,40^ the role of TREM1 in psoriasis is still a subject of debate. TREM1-*ko* mice exhibited responses to imiquimod similar to TREM1 positive mice. ^41^ In PN skin, the ‘IL-13 signaling’ pathway was enriched. Circulating IL-13 levels have been found to be higher in psoriatic cases compared to healthy controls, however, no correlation was established between IL-13 and PASI scores. ^42^ Additionally, the ‘Degradation of α-tocopherol’ pathway was enriched. Reduced levels of this antioxidant have been reported in psoriasis, and is suggested to promote atherosclerosis, a known comorbidity in psoriasis. ^43^

A major strength of this study is the large number of samples included, increasing the robustness of the results. However, for the severity specific analysis, only local samples were utilized, resulting in reduced power for this analysis. We validated the DEGs from the local dataset by correlation with the other datasets which overall confirmed strong correlations between the datasets. The California dataset was less correlated with the other datasets included, which may reflect differences in disease severity, given that the cases included in this study had higher PASI scores. Additionally, the psoriatic samples in the California study were collected from individuals who qualified for biological treatment. Differences in anatomical location of the samples might have also influences these correlations, as the NN skin was collected from surplus skin from plastic surgery procedures. ^44,45^

Functional follow-up studies would be needed to ascertain whether the observed changes in gene expression are casual or consequential in relation to psoriasis and/or increasing psoriasis severity. Moreover, all samples in the local datasets were from European ancestry individuals, which may limit the generalizability of our findings. To the best of our knowledge, this study represents the largest RNA-seq investigation of psoriatic skin to date, enabling a robust characterization of the psoriasis transcriptome. In summary, our findings reveal significant IL-17 effects even in non-lesional psoriatic skin, thereby providing further support for the pro-inflammatory signature of psoriasis. Furthermore, we have identified a comprehensive mRNA-based signature of psoriasis severity. We propose that these DEGs could serve as promising candidates for future studies aimed at developing biomarkers to predict disease trajectories and severity in psoriasis.

## Acknowledgements

We are grateful to all the volunteers who contributed to our study as well as to assistance from staff at the Clinical Research Facility, St. Olavs hospital, Trondheim University Hospital, Norway, Biobank1, Trondheim, Norway, the Clinical Research Unit, University Hospital of North Norway, Norway, the Research Biobank, University Hospital of North Norway, Norway and the Tromsø Study, UiT The Arctic University of Norway, Norway. The total RNA extraction, library preparation and RNA sequencing were performed in close collaboration with the Genomics Core Facility (GCF), at the Norwegian University of Science and Technology, NTNU, Trondheim, Norway. The GCF is funded by the Faculty of Medicine and Health Sciences at NTNU and Central Norway Regional Health Authority. All analyses were performed using digital laboratories in HUNT Cloud at the Norwegian University of Science and Technology, NTNU, Trondheim, Norway. We are grateful for outstanding support from the HUNT Cloud community. This work was supported by the Liaison Committee for Education, Research and Innovation in Central Norway (Grant number: 90154700) and the Joint Research Committee between St. Olavs hospital and the Faculty of Medicine and Health Sciences, NTNU (Grant numbers: 46055600-101 and 90053000). ML, PS, KH, KC, and LCO and SAH work in a research unit funded by Stiftelsen Kristian Gerhard Jebsen (Grant number: SKGJ-MED-015); Faculty of Medicine and Health Sciences, NTNU; The Liaison Committee for Education, Research and Innovation in Central Norway; and the Joint Research Committee between St. Olavs hospital and the Faculty of Medicine and Health Sciences, NTNU. ML was supported by a research grant from the Liaison Committee for education, research and innovation in Central Norway. Marita Jenssen, Kjersti Danielsen and Anne-Sofie Furberg were supported by grants from the Northern Norway Regional Health Authority (grant number HNF1361-17 [MJ] and SFP1167-14 [KD, ASF]), the Odd Berg Medical Research Foundation (MJ, KD) and the Norwegian Society of Dermatology and Venereology (MJ). The funders had no influence on study design, data collection and analysis, decision to publish, or preparation of the manuscript.

## Data availability statement

The normalized count matrix of 22 530 expressed genes from the local dataset is available from the corresponding authors upon reasonable request. Upon publication, count data will be deposited at NCBI GEO.

