## Supplementary materials for "Meta-analysis of RNA sequencing data from 534 skin samples shows substantial IL-17 effects in non-lesional psoriatic skin"

Supplementary table 1. Distribution of DEG biotypes in the meta-analysis for PP/NN, PP/PN and PN/NN.

|  | **PP/NN** | | **PP/PN** | | **PN/NN** | |
| --- | --- | --- | --- | --- | --- | --- |
| **biotype** | **Frequency** | **%** | **Frequency** | **%** | **Frequency** | **%** |
| IG_C_gene | 1 | 0.04 | 2 | 0.1 | 1 | 1.72 |
| TEC | 16 | 0.71 | 14 | 0.69 | 2 | 3.45 |
| TR_C_gene | 2 | 0.09 | 2 | 0.1 | 0 | 0 |
| antisense | 114 | 5.02 | 88 | 4.36 | 1 | 1.72 |
| bidirectional_promoter_lncRNA | 1 | 0.04 | 0 | 0 | 0 | 0 |
| lincRNA | 81 | 3.57 | 65 | 3.22 | 2 | 3.45 |
| misc_RNA | 1 | 0.04 | 0 | 0 | 0 | 0 |
| polymorphic_pseudogene | 1 | 0.04 | 1 | 0.05 | 0 | 0 |
| processed_pseudogene | 13 | 0.57 | 10 | 0.5 | 1 | 1.72 |
| processed_transcript | 23 | 1.01 | 19 | 0.94 | 0 | 0 |
| protein_coding | 1 970 | 86.82 | 1 776 | 87.96 | 50 | 86.21 |
| sense_intronic | 4 | 0.18 | 2 | 0.1 | 0 | 0 |
| sense_overlapping | 3 | 0.13 | 1 | 0.05 | 0 | 0 |
| transcribed_processed_pseudogene | 4 | 0.18 | 2 | 0.1 | 0 | 0 |
| transcribed_unitary_pseudogene | 4 | 0.18 | 2 | 0.1 | 0 | 0 |
| transcribed_unprocessed_pseudogene | 21 | 0.93 | 24 | 1.19 | 0 | 0 |
| unprocessed_pseudogene | 10 | 0.44 | 10 | 0.5 | 1 | 1.72 |
| snoRNA | 0 | 0 | 1 | 0.05 | 0 | 0 |
| **Total** | **2 269** | **100** | **2 019** | **100** | **58** | **100** |

DEGs defined as |log_2_ FC| >1 and FDR <0.1 in the given contrast. RNA species categorized into four categories as follows; i. protein coding: ´protein_coding´, ii. long non-coding: ´3prime_overlapping_ncRNA´, ´antisense´, ´bidirectional_promoter_lncRNA´, ´lincRNA´, ´macro_lncRNA´, ´sense_intronic´, ´sense_overlapping´, iii. pseudogene: ´IG_C_pseudogene´, ´IG_J_pseudogene´, ´IG_V_pseudogene´, ´IG_pseudogene´, ´TR_J_pseudogene´, ´TR_V_pseudogene´, ´polymorphic_pseudogene´, ´processed_pseudogene´, ´pseudogene´, ´transcribed_processed_pseudogene´, ´transcribed_unitary_pseudogene´, ´transcribed_unprocessed_pseudogene´, ´translated_processed_pseudogene´, ´unitary_pseudogene´, ´unprocessed_pseudogene´, iv. other: ‘IG_C_gene’, ‘IG_D_gene’, ´IG_V_gene´, ´Mt_rRNA´, ´Mt_tRNA´, ´TEC´, ´TR_C_gene´, ´TR_D_gene´, ´TR_J_gene´, ´TR_V_gene´, ´miRNA´, ´misc_RNA´, ´non_coding´, ´processed_transcript´, ´rRNA´, ´ribozyme´, ´sRNA´, ´scRNA´, ´scaRNA´, ´snRNA´, ´snoRNA´, ´vaultRNA´, ´IG_J_gene´.

Supplementary table 2. Datasets and their corresponding number of PP, PN and NN samples included in the local dataset and meta-analysis.

| Data source | NCBI  GEO accession number | Number of samples | | | Reference |
| --- | --- | --- | --- | --- | --- |
|  |  | PP | PN | NN |  |
| NTNU, Trondheim, Norway |  | 71 | 74 | 46 | Solvin et al. 2021 |
| Ann Arbor, MI, USA | Superseries GSE63980 (GSE54456 and GSE63979) | 92 | 0 | 82 | Tsoi et al. 2015, Li et al. 2014 |
|  |  | 7 | 27 | 8 | Greenberg et al. 2020 |
| Kiel, Germany | GSE121212 | 28 | 27 | 38 | Tsoi et al. 2019 |
| San Francisco, CA, USA | GSE74697 | 18 | 0 | 16 | Gupta et al. 2016, Ahn et al. 2016 |
| Total |  | 216 | 128 | 190 | 534 |

Supplementary table 3. Number of differentially expressed genes (FDR ≤0.1 and |log_2_ FC| ≥1) identified in the local dataset and meta-analysed datasets for each comparison pair.

|  | Local dataset | | | Meta-analysis | | |
| --- | --- | --- | --- | --- | --- | --- |
|  | PP/NN | PP/PN | PN/NN | PP/NN | PP/PN | PN/NN |
| Upregulated | 2584 | 2441 | 20 | 1115 | 1005 | 44 |
| Downregulated | 2408 | 2267 | 9 | 1154 | 1014 | 14 |
| Total | 4992 | 4708 | 29 | 2269 | 2019 | 58 |

Supplementary table 4. Number and proportion (%) of differentially expressed genes (FDR ≤0.1 and |log_2_ FC| ≥1) across different RNA biotypes in the meta-analysis for each comparison pair.

|  | Protein-coding | Long non-coding RNA | Pseudogene | Other |
| --- | --- | --- | --- | --- |
| PP/NN, *n* (%)  (N= 2 269) | 1 970 (86.8) | 203 (9.0) | 53 (2.3) | 43 (1.9) |
| PP/PN, *n* (%)  (N= 2 019) | 1 776 (88.0) | 156 (7.7) | 49 (2.4) | 38 (1.9) |
| PN/NN, *n* (%)  (N = 58) | 50 (86.2) | 3 (5.2) | 2 (3.5) | 3 (5.2) |

Supplementary table 5. List of top 10 DEGs sorted by absolute FC (|log_2_ FC|) for the different biotypes (long non-coding, pseudogene and other) in PP/NN of the meta-analysis.

| **Long non-coding RNA** | | | | | | |
| --- | --- | --- | --- | --- | --- | --- |
| genes | hgnc_symbol | logfc | AveExpr | PValue | adjpval | gene_name |
| ENSG00000260673 |  | 4.9090468 | -1.0919437 | 1.15E-144 | 2.79E-142 | AL034376.2 |
| ENSG00000272666 | KLHDC7B-DT | 4.3487579 | -0.6955428 | 3.23E-127 | 4.05E-125 | U62317.2 |
| ENSG00000271856 | LINC01215 | 3.8687969 | -1.6692431 | 6.05E-102 | 3.50E-100 | LINC01215 |
| ENSG00000268416 | | -3.7562113 | 0.61111433 | 7.80E-127 | 9.50E-125 | AC010329.1 |
| ENSG00000253417 | LINC02159 | 3.7427216 | -0.6238631 | 5.13E-89 | 1.95E-87 | LINC02159 |
| ENSG00000253745 | | -3.6920233 | -2.0274027 | 3.60E-54 | 4.51E-53 | AC091163.1 |
| ENSG00000253161 | LINC01605 | 3.5855339 | 0.42182954 | 4.40E-109 | 3.09E-107 | LINC01605 |
| ENSG00000231971 | CT69 | 3.3683749 | -1.2108064 | 1.05E-90 | 4.18E-89 | AL078590.2 |
| ENSG00000257700 | | -3.3127338 | -0.5703012 | 4.01E-76 | 1.04E-74 | AC055716.3 |
| ENSG00000254027 | LNMICC | 3.3003517 | 1.0368931 | 1.52E-43 | 1.32E-42 | AC009902.2 |
| **Pseudogene** | | | | | | |
| genes | hgnc_symbol | logfc | AveExpr | PValue | adjpval | gene_name |
| ENSG00000229035 | SPRR2C | 6.9674807 | -0.5972195 | 1.66E-139 | 2.96E-137 | SPRR2C |
| ENSG00000255128 | HSPD1P3 | 4.6851755 | -2.7821418 | 5.32E-86 | 1.85E-84 | HSPD1P3 |
| ENSG00000232022 | FAAHP1 | 3.633708 | -1.5786548 | 1.89E-98 | 9.45E-97 | FAAHP1 |
| ENSG00000214822 | KRT16P3 | 3.3355384 | -1.8722039 | 1.35E-63 | 2.29E-62 | KRT16P3 |
| ENSG00000214856 | KRT16P1 | 3.1657232 | -2.1844902 | 3.33E-60 | 5.05E-59 | KRT16P1 |
| ENSG00000227300 | KRT16P2 | 3.0194922 | -0.6768563 | 4.47E-47 | 4.46E-46 | KRT16P2 |
| ENSG00000160766 | GBAP1 | 2.8039553 | 1.7118835 | 3.15E-113 | 2.55E-111 | GBAP1 |
| ENSG00000268100 | ZNF725P | -2.773317 | -2.4574528 | 2.58E-37 | 1.82E-36 | ZNF725P |
| ENSG00000159247 | TUBBP5 | 2.6047894 | -0.5933434 | 1.11E-42 | 9.39E-42 | TUBBP5 |
| ENSG00000234964 | FABP5P7 | 2.4685311 | -0.8166179 | 5.53E-104 | 3.42E-102 | FABP5P7 |
| **Other** | | | | | | |
| genes | hgnc_symbol | logfc | AveExpr | PValue | adjpval | gene_name |
| ENSG00000279460 |  | -3.6889494 | -1.7951569 | 3.20E-52 | 3.77E-51 | AC112243.1 |
| ENSG00000237943 | PRKCQ-AS1 | 3.1389291 | -1.0478827 | 4.51E-67 | 8.62E-66 | PRKCQ-AS1 |
| ENSG00000280032 |  | 2.9780619 | -0.2206177 | 1.73E-99 | 8.96E-98 | AP002800.1 |
| ENSG00000261606 |  | 2.3430939 | -2.3693285 | 3.45E-60 | 5.23E-59 | AC091230.1 |
| ENSG00000231290 | APCDD1L-DT | -2.3331971 | -1.2177667 | 6.40E-48 | 6.56E-47 | APCDD1L-AS1 |
| ENSG00000236333 | TRHDE-AS1 | -2.3288323 | -0.107277 | 1.67E-37 | 1.19E-36 | TRHDE-AS1 |
| ENSG00000276170 | 2.2888924 | 1.0084775 | 5.97E-58 | 8.50E-57 | AC244153.1 |  |
| ENSG00000279296 | PRAL | 2.2695469 | -0.7182502 | 4.90E-45 | 4.51E-44 | PRAL |
| ENSG00000279693 |  | 2.2133619 | 1.0586958 | 8.84E-71 | 1.94E-69 | AC099521.2 |
| ENSG00000130600 | H19 | -2.1931934 | 2.4971021 | 1.04E-42 | 8.82E-42 | H19 |

Supplementary table 6. List of top 10 DEGs sorted by absolute FC (|log_2_ FC|) for the different biotypes (long non-coding, pseudogene and other) in PN/NN of the meta-analysis.

| **Long non-coding RNA** | | | | | | |
| --- | --- | --- | --- | --- | --- | --- |
| genes | hgnc_symbol | logfc | AveExpr | PValue | adjpval | gene_name |
| ENSG00000260673 |  | 1.3348557 | -1.0919437 | 1.69E-11 | 2.30E-08 | AL034376.2 |
| ENSG00000228318 |  | 1.1104708 | -0.8279738 | 2.37E-08 | 9.31E-06 | AP001610.1 |
| ENSG00000253745 |  | -1.1039468 | -2.0274027 | 0.00005756 | 0.00271538 | AC091163.1 |
| **Pseudogene** | | | | | | |
| genes | hgnc_symbol | logfc | AveExpr | PValue | adjpval | gene_name |
| ENSG00000229035 | SPRR2C | 1.9589144 | -0.5972195 | 5.90E-12 | 1.16E-08 | SPRR2C |
| ENSG00000255128 | HSPD1P3 | 1.9547197 | -2.7821418 | 3.66E-13 | 9.24E-10 | HSPD1P3 |
| **Other** | | | | | | |
| genes | hgnc_symbol | logfc | AveExpr | PValue | adjpval | gene_name |
| ENSG00000279296 | PRAL | 1.0332221 | -0.7182502 | 1.21E-07 | 0.00002971 | PRAL |
| ENSG00000279460 |  | -1.029307 | -1.7951569 | 0.00023442 | 0.00673682 | AC112243.1 |
| ENSG00000211675 | IGLC1 | 1.0203205 | -2.9782076 | 1.14E-07 | 0.00002858 | IGLC1 |

Supplementary table 7. List of top 10 DEGs sorted by absolute FC (|log_2_ FC|) for the different biotypes (long non-coding, pseudogene and other) in PP skin of the severity-specific analysis.

| **Long non-coding RNA** | | | | | |
| --- | --- | --- | --- | --- | --- |
| none |  |  |  |  |  |
| **Pseudogene** | | | | | |
| genes | logfc | AveExpr | PValue | adjpval | gene_name |
| ENSG00000224689 | 3.2928483 | -4.413253 | 7.34E-07 | 0.01653659 | ZNF812P |
| **Other** | | | | | |
| none |  |  |  |  |  |

Supplementary table 8. List of top 10 DEGs sorted by absolute FC (|log_2_ FC|) for the different biotypes (long non-coding, pseudogene and other) in PN skin of the severity-specific analysis.

| **Long non-coding RNA** | | | | | |
| --- | --- | --- | --- | --- | --- |
| genes | logfc | AveExpr | PValue | adjpval | gene_name |
| ENSG00000282851 | -3.3349754 | 1.3357812 | 0.00017606 | 0.07201846 | BISPR |
| ENSG00000241912 | 2.8321953 | -4.4921473 | 0.00016921 | 0.07123064 | AC078788.2 |
| ENSG00000266126 | 2.6197336 | -2.4078737 | 0.00010863 | 0.06478057 | AC005730.3 |
| ENSG00000259495 | -2.5956093 | -2.122754 | 0.00035434 | 0.0986562 | AC016705.2 |
| ENSG00000278385 | 2.5264644 | -4.4798304 | 0.00005788 | 0.042067 | AC121338.2 |
| ENSG00000124915 | 2.4778531 | -4.4038592 | 0.0001094 | 0.06478057 | AP002380.1 |
| ENSG00000223342 | 2.3116842 | -5.2446527 | 0.00011365 | 0.06478057 | AL158817.1 |
| ENSG00000273958 | 2.248539 | -1.6167916 | 0.0001519 | 0.06921137 | AC009560.1 |
| ENSG00000216863 | 2.2099806 | -5.469427 | 0.00004355 | 0.03634413 | LY86-AS1 |
| **Pseudogene** | | | | | |
| genes | logfc | AveExpr | Pvalue | adjpval | gene_name |
| ENSG00000251595 | -2.7806706 | -0.6188293 | 0.0000259 | 0.03391707 | ABCA11P |
| ENSG00000215105 | -2.4163466 | -1.1273385 | 0.00024529 | 0.07797654 | TTC3P1 |
| ENSG00000233609 | 2.3567615 | -5.3238905 | 0.00004099 | 0.03634413 | RPL10P19 |
| ENSG00000262381 | 2.2399573 | -0.9155493 | 0.00034475 | 0.0986562 | MTATP6P24 |
| **Other** | | | | | |
| genes | logfc | AveExpr | Pvalue | adjpval | gene_name |
| ENSG00000254929 | -2.8511956 | -0.6221567 | 4.70E-06 | 0.01764423 | AL591684.2 |

Supplementary table 9. Top 20 enriched canonical pathways identified in PP/NN of the meta-analysis.

| Ingenuity Canonical Pathways | -log(p-value) | Ratio | z-score |
| --- | --- | --- | --- |
| Pathogen Induced Cytokine Storm Signaling Pathway | 14.5 | 0.216 | 5.143 |
| Agranulocyte Adhesion and Diapedesis | 14.4 | 0.267 | NA |
| S100 Family Signaling Pathway | 13.2 | 0.166 | 1.591 |
| Granulocyte Adhesion and Diapedesis | 12.8 | 0.265 | NA |
| Atherosclerosis Signaling | 11.7 | 0.293 | NA |
| LPS/IL-1 Mediated Inhibition of RXR Function | 8.36 | 0.201 | 3 |
| Role of Hypercytokinemia/hyperchemokinemia in the Pathogenesis of Influenza | 8.32 | 0.302 | 4.707 |
| Kinetochore Metaphase Signaling Pathway | 7.6 | 0.261 | 3.53 |
| LXR/RXR Activation | 7.1 | 0.244 | -2.746 |
| Glucocorticoid Receptor Signaling | 6.69 | 0.148 | NA |
| Phagosome Formation | 6.02 | 0.138 | 0.612 |
| Calcium Signaling | 5.96 | 0.186 | -1.177 |
| Role Of Osteoblasts In Rheumatoid Arthritis Signaling Pathway | 5.94 | 0.18 | 1.508 |
| CREB Signaling in Neurons | 5.91 | 0.142 | -1.427 |
| Dilated Cardiomyopathy Signaling Pathway | 5.63 | 0.207 | 1.877 |
| Breast Cancer Regulation by Stathmin1 | 5.49 | 0.14 | -0.663 |
| MSP-RON Signaling Pathway | 5.46 | 0.293 | NA |
| Airway Pathology in Chronic Obstructive Pulmonary Disease | 5.36 | 0.22 | NA |
| Hepatic Fibrosis / Hepatic Stellate Cell Activation | 5.27 | 0.186 | NA |

Supplementary table 10. Top 20 predicted upstream regulators in PP/NN of the meta-analysis.

| Upstream Regulator | Expr Log Ratio | Molecule Type | Predicted Activation State | Activation z-score | p-value of overlap |
| --- | --- | --- | --- | --- | --- |
| TNF | 1.07 | cytokine | Activated | 10.684 | 5.19E-58 |
| IFNG |  | cytokine | Activated | 9.281 | 2.77E-56 |
| beta-estradiol |  | chemical - endogenous mammalian |  | 1.063 | 6.97E-49 |
| dexamethasone |  | chemical drug | Inhibited | -5.977 | 7.85E-47 |
| STAT3 | 1.12 | transcription regulator | Activated | 3.446 | 3.12E-46 |
| lipopolysaccharide |  | chemical drug | Activated | 11.482 | 6.08E-45 |
| Interferon alpha |  | group | Activated | 8.804 | 9.45E-44 |
| IL1B | 2.641 | cytokine | Activated | 7.085 | 1.12E-41 |
| IL6 |  | cytokine | Activated | 4.983 | 3.24E-41 |
| tetradecanoylphorbol acetate |  | chemical drug | Activated | 5.315 | 6.7E-41 |
| Eldr |  | other | Activated | 6.751 | 2.69E-40 |
| TGFB1 | 0.151 | growth factor | Activated | 2.627 | 1.14E-38 |
| CEBPB | 0.451 | transcription regulator | Activated | 6.07 | 3.62E-37 |
| calcitriol |  | chemical drug | Inhibited | -2.103 | 1.85E-36 |
| Immunoglobulin |  | complex |  | 1.809 | 4.94E-36 |
| IL4 |  | cytokine |  | 1.873 | 2.15E-34 |
| STAT1 | 1.735 | transcription regulator | Activated | 6.508 | 2.27E-34 |
| OSM |  | cytokine | Activated | 7.18 | 6.62E-33 |
| tretinoin |  | chemical - endogenous mammalian | Activated | 2.619 | 2.28E-32 |

Supplementary table 11. Top 20 enriched canonical pathways in PN/NN of the meta-analysis.

| Ingenuity Canonical Pathways | -log(p-value) | Ratio | z-score |
| --- | --- | --- | --- |
| Role of IL-17A in Psoriasis | 12.6 | 0.429 | 2.449 |
| IL-17A Signaling in Gastric Cells | 4.62 | 0.115 | NA |
| S100 Family Signaling Pathway | 4.43 | 0.0117 | 3 |
| IL-17 Signaling | 4.28 | 0.0267 | 2.236 |
| Macrophage Classical Activation Signaling Pathway | 4.26 | 0.0265 | 2.236 |
| Granulocyte Adhesion and Diapedesis | 4.26 | 0.0265 | NA |
| Agranulocyte Adhesion and Diapedesis | 4.04 | 0.0238 | NA |
| LXR/RXR Activation | 3.83 | 0.0325 | NA |
| Osteoarthritis Pathway | 3.8 | 0.0212 | NA |
| Role of IL-17A in Arthritis | 3.59 | 0.0526 | NA |
| IL-17A Signaling in Airway Cells | 3.38 | 0.0448 | NA |
| Role of Hypercytokinemia/hyperchemokinemia in the Pathogenesis of Influenza | 3.06 | 0.0349 | NA |
| Pathogen Induced Cytokine Storm Signaling Pathway | 2.91 | 0.0135 | 2.236 |
| Airway Pathology in Chronic Obstructive Pulmonary Disease | 2.67 | 0.0254 | NA |
| Airway Inflammation in Asthma | 2.63 | 0.0606 | NA |
| Atherosclerosis Signaling | 2.52 | 0.0226 | NA |
| Role Of Chondrocytes In Rheumatoid Arthritis Signaling Pathway | 2.45 | 0.0213 | NA |
| Retinoate Biosynthesis I | 2.43 | 0.0476 | NA |
| Estrogen Biosynthesis | 2.37 | 0.0444 | NA |

Supplementary table 12. Top 20 predicted upstream regulators in PN/NN of the meta-analysis.

| Upstream Regulator | Expr Log Ratio | Molecule Type | Predicted Activation State | Activation z-score | p-value of overlap |
| --- | --- | --- | --- | --- | --- |
| IL1A | -0.384 | cytokine | Activated | 3.662 | 1.28E-16 |
| IL22 |  | cytokine | Activated | 2.778 | 1.92E-16 |
| IL17C |  | cytokine | Activated | 2.789 | 4.69E-16 |
| BCL3 | 0.126 | transcription regulator |  | 1.273 | 1.14E-14 |
| OSM |  | cytokine | Activated | 3.759 | 4.26E-13 |
| IL36A | 2.442 | cytokine | Activated | 2.618 | 7.69E-13 |
| IL17A |  | cytokine | Activated | 3.306 | 2.02E-12 |
| IL17R |  | complex | Activated | 2.425 | 6.31E-12 |
| NFKBIZ | 0.363 | transcription regulator | Activated | 2.415 | 2.94E-11 |
| EHF | 0.059 | transcription regulator | Activated | 2.646 | 1.24E-09 |
| IL22RA1 | -0.08 | transmembrane receptor |  |  | 2.18E-09 |
| HBEGF | -0.488 | growth factor |  | 1.67 | 2.66E-09 |
| peptidoglycan |  | chemical - endogenous non-mammalian | Activated | 2.351 | 3.44E-09 |
| SCD | 0.356 | enzyme |  | -1.414 | 5.68E-09 |
| ELF3 | -0.135 | transcription regulator |  | 1.985 | 1.61E-08 |
| TNIP1 | 0.017 | other | Inhibited | -2.219 | 2.39E-08 |
| WZB117 |  | chemical reagent |  |  | 3.51E-08 |
| IL1RL2 | 0.079 | transmembrane receptor |  | 1.982 | 6.59E-08 |
| TLR3 | -0.155 | transmembrane receptor | Activated | 2.758 | 8.32E-08 |

Supplementary table 13. Top enriched canonical pathways in PP skin of the severity specific-analysis.

| Ingenuity Canonical Pathways | -log(p-value) | Ratio | z-score |
| --- | --- | --- | --- |
| Role of IL-17A in Psoriasis | 2.45 | 0.0714 | NA |
| Differential Regulation of Cytokine Production in Intestinal Epithelial Cells by IL-17A and IL-17F | 2.23 | 0.0435 | NA |
| Coagulation System | 2.05 | 0.0286 | NA |
| IL-17A Signaling in Airway Cells | 1.77 | 0.0149 | NA |
| TREM1 Signaling | 1.71 | 0.013 | NA |
| Unfolded protein response | 1.64 | 0.0111 | NA |
| IL-13 Signaling Pathway | 1.53 | 0.00862 | NA |
| HMGB1 Signaling | 1.38 | 0.00599 | NA |
| Aldosterone Signaling in Epithelial Cells | 1.37 | 0.00581 | NA |
| IL-17 Signaling | 1.33 | 0.00535 | NA |
| NRF2-mediated Oxidative Stress Response | 1.23 | 0.00422 | NA |
| Protein Ubiquitination Pathway | 1.17 | 0.00365 | NA |
| Th17 Activation Pathway | 0.932 | 0.00207 | NA |
| Glucocorticoid Receptor Signaling | 0.854 | 0.00172 | NA |
| S100 Family Signaling Pathway | 0.742 | 0.0013 | NA |

Supplementary table 14. Top 20 predicted upstream regulators in PP skin of the severity-specific analysis.

| Upstream Regulator | Expr Log Ratio | Molecule Type | p-value of overlap |
| --- | --- | --- | --- |
| DACH1 | 0.551 | transcription regulator | 5.89E-05 |
| ganglioside GM2 |  | chemical - endogenous mammalian | 0.000283 |
| proadifen |  | chemical reagent | 0.000283 |
| 5,6-epoxyeicosatrienoic acid |  | chemical - endogenous mammalian | 0.000566 |
| defibrotide |  | biologic drug | 0.000849 |
| mir-340 |  | microRNA | 0.00113 |
| WZB117 |  | chemical reagent | 0.00113 |
| ganglioside GD1b |  | chemical - endogenous mammalian | 0.00142 |
| anthralin |  | chemical drug | 0.00142 |
| norethindrone acetate |  | chemical drug | 0.0017 |
| nicotine |  | chemical drug | 0.00191 |
| MEFV | 1.474 | other | 0.00198 |
| PDE4D | -0.042 | enzyme | 0.00198 |
| retroinverso ERG inhibitory peptide 2 |  | chemical reagent | 0.00198 |
| retroinverso ERG inhibitory peptide 1 |  | chemical reagent | 0.00198 |
| mercury |  | chemical toxicant | 0.00198 |
| ganglioside GD1a |  | chemical - endogenous mammalian | 0.00226 |
| ganglioside GM1 |  | chemical - endogenous mammalian | 0.00226 |
| MIA | -0.551 | other | 0.00226 |

Supplementary table 15. Top 20 enriched canonical pathways in PN skin of the severity-specific analysis.

| Ingenuity Canonical Pathways | -log(p-value) | Ratio | z-score |
| --- | --- | --- | --- |
| IL-13 Signaling Pathway | 1.94 | 0.0172 | NA |
| α-tocopherol Degradation | 1.86 | 0.1 | NA |
| Parkinson's Signaling | 1.65 | 0.0625 | NA |
| Glutaryl-CoA Degradation | 1.58 | 0.0526 | NA |
| IL-17 Signaling | 1.55 | 0.0107 | NA |
| ID1 Signaling Pathway | 1.49 | 0.00995 | NA |
| Tryptophan Degradation III (Eukaryotic) | 1.44 | 0.0385 | NA |
| Role Of Osteoblasts In Rheumatoid Arthritis Signaling Pathway | 1.34 | 0.0082 | NA |
| Thyroid Hormone Metabolism II (via Conjugation and/or Degradation) | 1.26 | 0.025 | NA |
| Oncostatin M Signaling | 1.23 | 0.0233 | NA |
| Spliceosomal Cycle | 1.18 | 0.0204 | NA |
| Nicotine Degradation III | 1.11 | 0.0172 | NA |
| PCP (Planar Cell Polarity) Pathway | 1.09 | 0.0167 | NA |
| Melatonin Degradation I | 1.08 | 0.0161 | NA |
| Nicotine Degradation II | 1.05 | 0.0152 | NA |
| Superpathway of Melatonin Degradation | 1.05 | 0.0149 | NA |
| Serotonin Degradation | 1.02 | 0.0141 | NA |
| ERK5 Signaling | 1.01 | 0.0135 | NA |
| Role of JAK family kinases in IL-6-type Cytokine Signaling | 0.979 | 0.0127 | NA |

Supplementary table 16. Top 20 predicted upstream regulators in PN skin of the severity-specific analysis.

| Upstream Regulator | Expr Log Ratio | Molecule Type | Predicted Activation State | Activation z-score | p-value of overlap |
| --- | --- | --- | --- | --- | --- |
| miR-629-5p (miRNAs w/seed GGGUUUA) |  | mature microRNA |  |  | 0.00142 |
| N-carbamyl-L-glutamate |  | chemical drug |  |  | 0.00142 |
| PRPF8 | 0.008 | other |  |  | 0.00283 |
| miR-637 (and other miRNAs w/seed CUGGGGG) |  | mature microRNA |  |  | 0.00283 |
| RANBP2 | 0.152 | enzyme |  |  | 0.00283 |
| 3,4-dihydroxybenzaldehyde |  | chemical - endogenous non-mammalian |  |  | 0.00424 |
| nomifensine |  | chemical drug |  |  | 0.00424 |
| chitosan |  | chemical - endogenous mammalian |  |  | 0.00424 |
| JAK1 | -0.079 | kinase |  |  | 0.00493 |
| MYOD1 |  | transcription regulator |  |  | 0.00544 |
| clomiphene |  | chemical drug |  |  | 0.00565 |
| fluoride |  | chemical - endogenous mammalian |  |  | 0.00617 |
| RUNX1 | 0.068 | transcription regulator |  |  | 0.00685 |
| advanced glycation end product 3 |  | chemical - endogenous mammalian |  |  | 0.00706 |
| DIPQUO |  | chemical reagent |  |  | 0.00706 |
| SLU7 | 0.17 | enzyme |  |  | 0.00706 |
| WBP11 | 0.128 | phosphatase |  |  | 0.00706 |
| ZBTB46 | -0.498 | transcription regulator |  |  | 0.00706 |
| EDIL3 | -0.569 | other |  |  | 0.00706 |

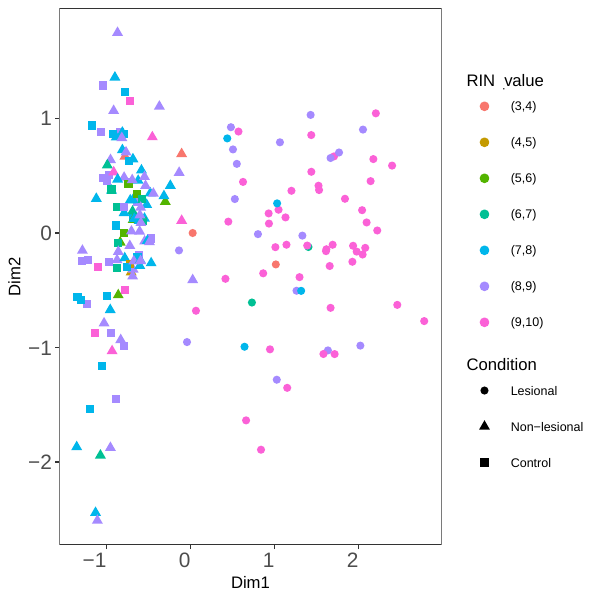

Supplementary figure 1. Multidimensional Scaling Plot (MDS) plot of samples in the local dataset by sample group and RIN value.
